# Cohort Profile: the Oxford Parkinson’s Disease Centre Discovery Cohort Magnetic Resonance Imaging sub-study (OPDC-MRI)

**DOI:** 10.1101/19005819

**Authors:** Ludovica Griffanti, Johannes C Klein, Konrad Szewczyk-Krolikowski, Ricarda A L Menke, Michal Rolinski, Thomas R Barber, Michael Lawton, Samuel G Evetts, Faye Begeti, Marie Crabbe, Jane Rumbold, Richard Wade-Martins, Michele T. Hu, Clare E Mackay

## Abstract

**Purpose:** The Oxford Parkinson’s Disease Centre (OPDC) Discovery Cohort magnetic resonance imaging (MRI) sub-study (OPDC-MRI) collects high quality multimodal brain MRI together with deep longitudinal clinical phenotyping in patients with Parkinson’s, at-risk individuals and healthy elderly participants. The primary aim is to detect pathological changes in brain structure and function, and develop, together with the clinical data, biomarkers to stratify, predict and chart progression in early-stage Parkinson’s and at-risk individuals.

**Participants:** Participants are recruited from the OPDC Discovery Cohort, a prospective, longitudinal study. Baseline MRI data is currently available for 290 participants: 119 patients with early idiopathic Parkinson’s, 15 Parkinson’s patients with pathogenic mutations of the LRRK2 or GBA genes, 68 healthy controls and 87 individuals at risk of Parkinson’s (asymptomatic carriers of GBA mutation and patients with idiopathic rapid eye movement sleep behaviour disorder - RBD).

**Findings to date:** Differences in brain structure in early Parkinson’s were found to be subtle, with small changes in the shape of the globus pallidus and evidence of alterations in microstructural integrity in the prefrontal cortex that correlated with performance on executive function tests. Brain function, as assayed with resting fMRI yielded more substantial differences, with basal ganglia connectivity reduced in early Parkinson’s, and RBD, but not Alzheimer’s, suggesting that the effect is pathology specific. Imaging of the substantia nigra with the more recent adoption of sequences sensitive to iron and neuromelanin content shows promising results in identifying early signs of Parkinsonian disease.

**Future plans:** Ongoing studies include the integration of multimodal MRI measures to improve discrimination power. Follow-up clinical data are now accumulating and will allow us to correlate baseline imaging measures to clinical disease progression. Follow-up MRI scanning started in 2015 and is currently ongoing, providing the opportunity for future longitudinal imaging analyses with parallel clinical phenotyping.

**Article Summary:** *Strengths and limitations of this study:*

- High quality 3T MRI data in a very well phenotyped and longitudinally followed cohort of Parkinson’s and RBD.
- All imaging data were acquired on the same MRI scanner, quite unique for a study of this duration. The protocol includes both standard sequences, comparable across other studies, and sequences acquired to investigate study-specific research questions.
- Clinical longitudinal data are acquired every 18 months and will be used to relate baseline imaging with clinical progression. Information about conversion to Parkinson’s of the at-risk individuals will also be available, providing the ultimate validation of potential biomarkers. MRI follow-up is also ongoing, which will allow longitudinal imaging analyses.
- Statistical maps of published results and support data relative to the analyses are available to share.
- OPDC-MRI phenotyping is deep and relatively frequent, however the size of the cohort is not at the level of population-level cohort studies. MRI sequences are high quality, but could not exploit the latest advances in the field in order to maintain continuity.

## Introduction

The Oxford Parkinson’s Disease Centre (OPDC)^1^ is a multidisciplinary research centre at the University of Oxford supported by Parkinson’s UK with funds from The Monument Trust. It was established in 2010 and brings together world-leaders in clinical neurology, neuroepidemiology, neuroimaging, proteomics, genomics, molecular genetics, transgenic Parkinson’s models, neuropharmacology, neurophysiology and neuropathology.

The centre was formed to understand the earliest events in the development of Parkinson’s, ultimately with a view to identifying the changes that occur before motor symptoms become apparent.

The overarching goals of the OPDC are to:

- Understand the progression of Parkinson’s
- Predict the onset of Parkinson’s
- Identify potential drug targets for Parkinson’s
- Develop new treatments that will prevent the development of Parkinson’s in at-risk individuals.

To these aims, the research activity is structured around three overlapping themes: 1. improved clinical cohorts for development of novel biomarkers; 2. improved cellular and genetic models of Parkinson’s pathologies and pathways; 3. novel animal models of early neuronal dysfunction in Parkinson’s.

Within theme 1, the OPDC Discovery Cohort is one of the largest and best-characterised cohorts of people with early motor-manifest and prodromal Parkinson’s in the world^2-4^. It is a prospective, longitudinal study that has recruited patients with early idiopathic Parkinson’s, healthy controls and individuals at risk of Parkinson’s.

The OPDC Discovery Cohort is designed by and for patients and is closely linked with the Parkinson’s UK local support group. Patient representatives are also involved in the funding/renewal and strategic oversight processes.

The aim of the OPDC Discovery Cohort is to provide a wealth of data to better understand the biology of premotor and early Parkinson’s, and to identify predictors of disease onset and progression. In addition to standardised assessments of motor and non-motor function, there is a particular interest in validating cutting-edge technologies to stratify, predict and chart progression in Parkinson’s, including brain imaging, saccadometry, smart-phone and wearable assessments.

The subset of the OPDC Discovery Cohort that underwent brain magnetic resonance imaging (MRI) constitutes the OPDC Discovery MRI (OPDC-MRI) sub-study and is the focus of this paper. Driven by the emerging evidence of novel imaging markers with high predictive value, like the study by Vaillancourt and colleagues^5^ and others mentioned in ^6^, this project started with the collection of a small number of scans. Thanks to the promising results and to accumulating evidence in the field about the potential of MRI to serve as a biomarker for manifest (e.g.^7 8^) and pre-motor Parkinson’s^9^, the imaging sub-study was further expanded, with the aim to develop biomarkers derived from multimodal MRI in order to:

- Detect damage and changes in brain structure and function in early-stage Parkinson’s and prodromal ‘at-risk’ individuals.
- Predict disease progression and understand its neural correlates.
- Stratify at-risk individuals to identify potential candidates for clinical trials.

In this cohort profile we will describe the cohort composition, the MRI data collected, and the processing pipelines that we developed. We will then report findings to date and illustrate our future plans for the cohort.

## Cohort description

### Eligibility criteria and recruitment

Participants of the OPDC-MRI sub-study were recruited from the OPDC Discovery cohort since 2010. Neurologists, Parkinson’s nurses, geriatricians and GPs from participating hospitals in the Thames Valley area (total population 2.1 million) were asked to identify all idiopathic Parkinson’s cases who were diagnosed by a neurologist (or a geriatrician with a specialist interest in Parkinson’s) within the previous three years, according to the UK PD Society Brain Bank Criteria for clinically probable idiopathic Parkinson’s disease^10^. All participating clinicians are regularly contacted to ensure screening of incident cases diagnosed since study onset. Eligible cases were approached by post and asked to contact the OPDC if interested in taking part in the study. Exclusion criteria for participation are: non-idiopathic parkinsonism, secondary parkinsonism due to head trauma or medication use, cognitive impairment precluding informed consent, dementia preceding motoric Parkinson’s by one year suggestive of dementia with Lewy bodies, or other features of atypical parkinsonism syndromes such as multiple system atrophy, progressive supranuclear palsy, corticobasal degeneration.

More details on the recruitment process, assessment and exclusion criteria for the OPDC Discovery Cohort are described elsewhere^2 11^. For the MRI sub-study, we aimed to scan Parkinson’s patients as quickly as possible after enrolment (within 3 years of diagnosis), but due to logistics and patient unavailabilities, scanning was performed up to 6 years after diagnosis (see Table 1 for details). Parkinson’s patients with more than mild head tremor or presence of dyskinesia/dystonia were excluded in order to minimise movement artefacts during imaging. The Parkinson’s imaging cohort includes sporadic patients (iPD, N=119) and patients with known pathogenic mutations of the Leucine-rich repeat kinase 2 (LRRK2; G2019S and R1441C; N=5) or glucocerebrosidase (GBA; L444P and N370S; N=10) genes. Please see ^4^ for further details on GBA and LRRK2 genotyping methods.

**Table 1.** Demographic and clinical characteristics of the OPDC-MRI cohort.

|  | iPD | PD-LRRK2 | PD-GBA | RBD | RBD-GBA | aGBA | HC |
| --- | --- | --- | --- | --- | --- | --- | --- |
| N | 119 | 5 | 10 | 74 | 3 | 8 | 68 |
| Age [years]<br>(mean±stdev) | 64.1<br>±10.1 | 66.0<br>±11.6 | 63.8<br>±10.3 | 65.8<br>±7.6 | 61.2<br>±7.4 | 65.9<br>±7.6 | 65.9<br>±8.7 |
| Gender [M/F] | 76/43 | 2/3 | 6/4 | 68/6 | 3/0 | 3/5 | 45/23 |
| Parkinson's disease<br>duration at time of<br>MRI [years]<br>(mean±stdev) | (n=117)<br>2.31<br>±1.52 | 3.06<br>±3.49 | 3.29<br>±1.81 | - | - | - | - |
| Time between MRI<br>and closest clinical<br>assessment [days]<br>(mean±stdev) | (n=118)<br>108<br>±104 | 79 ± 105 | 140<br>±107 | 115 ± 90 | 6 ± 4 | (n=6)<br>273 ±25 | (n=64)<br>387±644 |
| Levodopa<br>equivalent dose<br>(LEDD)<br>(mean±stdev) § | (n=117)<br>335±243 | 340<br>±198 | 463<br>±243 | - | - | - | - |
| Hoehn & Yahr<br>(mean±stdev) § | (n=118)<br>1.79±0.5<br>7 | 2.40<br>±0.89 | 2.10<br>±0.88 | (n=71)<br>0.03<br>±0.17 | 0 | (n=6) 0 | (n=64) 0 |
| UPDRS III<br>(mean±stdev) § # | (n=118)<br>24.0<br>±10.4 | 41.2<br>±17.8 | 28.4<br>±14.9 | (n=74)<br>4.5 ±4.0 | 3.0 ±1.7 | (n=6)<br>3.3 ±4.2 | (n=64)<br>1.8 ±2.5 |
| MoCA<br>(mean±stdev) § ## | (n=117)<br>26.4<br>±2.7 | 26.0<br>±2.2 | 24.4<br>±3.1 | (n=73)<br>25.6<br>±2.7 | (n=2)<br>26.0<br>±0.0 | (n=6)<br>26.7<br>±1.9 | (n=62)<br>27.5<br>±2.0 |

When data is missing for some participants, the number of values available is specified in brackets.
§ = Evaluated at closest clinical assessment (controls only have a baseline clinic visit at enrolment).

The healthy control group is comprised of 68 participants with no family history of parkinsonism, many of whom were spouses and friends of Parkinson’s participants. Healthy controls were not receiving any medications known to affect the dopaminergic system. The at-risk group includes 74 patients with idiopathic rapid eye movement (REM) sleep behavior disorder (RBD), diagnosed with polysomnography according to International Classification of Sleep Disorders criteria^12^ (for more details see ^4^), RBD patients with a pathogenic mutation of the GBA gene (N=3), and asymptomatic carriers of GBA gene pathogenic mutations (N=8).

Additional exclusion criteria for the MRI sub-study for all groups were contraindications to MRI scanning, including a history of claustrophobia, incompatible metal foreign body or suspicion of such, unresolved metallic injury to the eye, or inability to travel to Oxford without assistance.

The study was undertaken with the understanding and written consent of each subject, with the approval of the local NHS ethics committee, and in compliance with national legislation and the Declaration of Helsinki.

### Data collection

Baseline MRI data were collected between November 2010 and December 2018. Follow-up data acquisition started in 2015 and is currently ongoing (see future plans section). Participants with Parkinson’s were scanned in a clinically defined “off” state, a minimum of 12h after the withdrawal of their dopaminergic medications.

#### Clinical data

Participants receive extensive assessment in designated research clinics as part of their participation in the OPDC Discovery cohort. The assessment, performed by a nurse and neurologist, includes a structured general medical interview, detailed characterisation of motor and non-motor features, and cognitive assessment (see ^2^ for details). Patients are followed up clinically every 18 months, while controls only have a baseline clinic visit. In a research clinic, we cannot formally diagnose patients who convert from RBD to Parkinson’s (or another neurodegenerative disorder). However, where history and examination suggest conversion to neurodegenerative disease, we alert the treating clinicians who then establish the diagnosis and ensure clinical management is in place. On the day of scanning, an additional UPDRS III assessment was performed (“off” in Parkinson’s). Table 1 summarises the main demographic and clinical characteristics of the OPDC-MRI sub-study cohort.

#### Imaging data

Scanning was performed at the Oxford Centre for Clinical Magnetic Resonance Research (OCMR) using a 3T Siemens Trio MRI scanner (Siemens, Erlangen, Germany) equipped with a 12-channel receive-only head coil. The neuroimaging protocol includes both structural and functional sequences and lasts approximately 45-50 minutes.

Within the allocated time, five modalities were always acquired (*core sequences*), while the remaining time was used to experiment with novel sequences, which changed during the study. We report here the details of two of them, introduced at a later stage based on new information becoming available in the field, and which showed promising preliminary results. We describe the other additional sequences we experimented in the supplementary material.

For each modality, pipelines for preprocessing and analysis of MRI data were developed and applied to the data to extract single-subject imaging variables (both summary and voxel-wise measurements). Here below we describe for each modality the rationale for acquisition, as well as the main preprocessing steps performed on the images to derive the measures of interest.

The main acquisition parameters and the number of available scans per group for each of the *core sequences* are listed in Table 2.

**Table 2.**
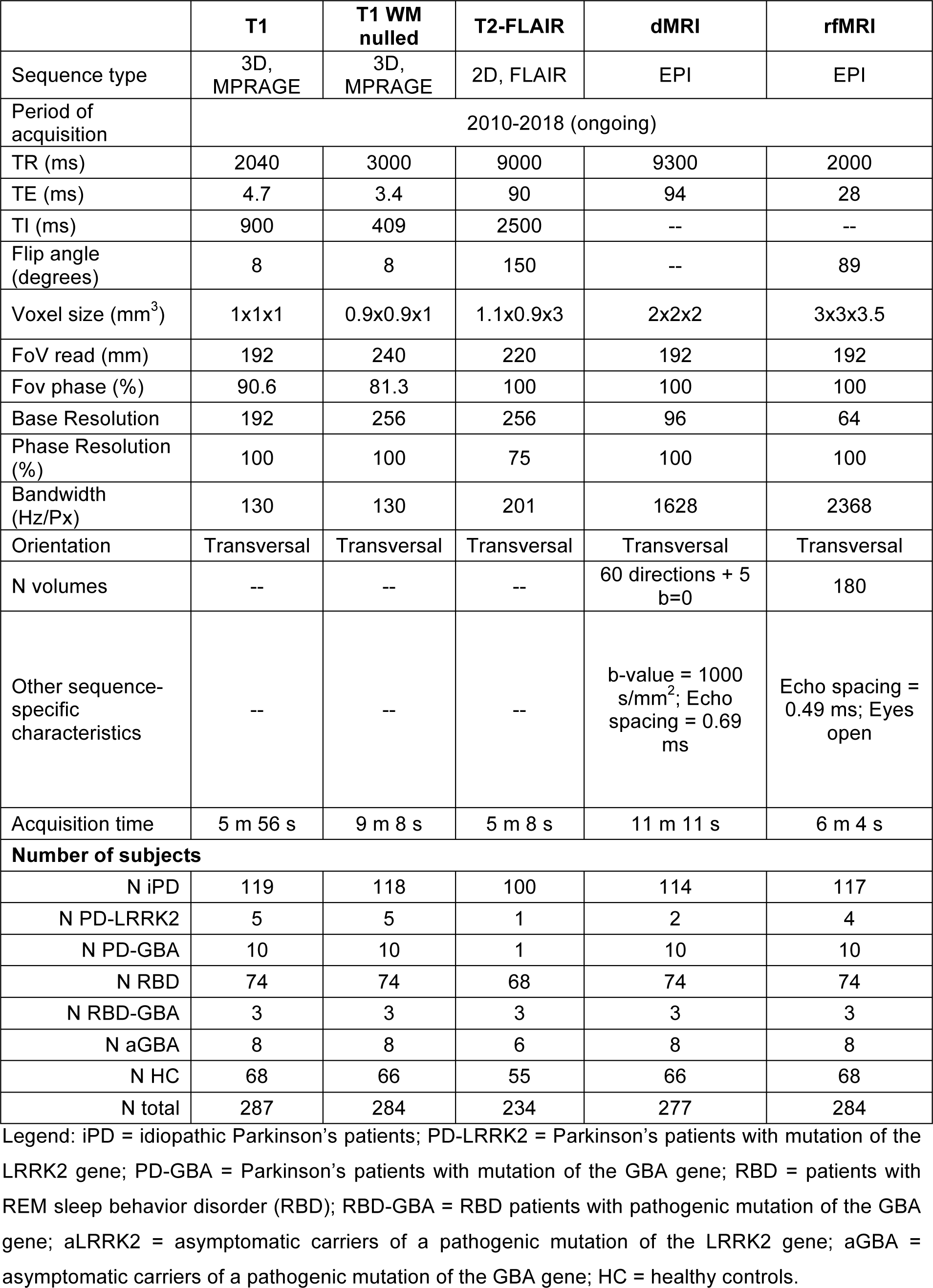
MRI core-sequences: parameters used in the study and number of available datasets for each modality.

|  | **T1** | **T1 WM nulled** | **T2-FLAIR** | **dMRI** | **rfMRI** |
| --- | --- | --- | --- | --- | --- |
| Sequence type | 3D, MPRAGE | 3D, MPRAGE | 2D, FLAIR | EPI | EPI |
| Period of acquisition | 2010-2018 (ongoing) |  |  |  |  |
| TR (ms) | 2040 | 3000 | 9000 | 9300 | 2000 |
| TE (ms) | 4.7 | 3.4 | 90 | 94 | 28 |
| TI (ms) | 900 | 409 | 2500 | -- | -- |
| Flip angle (degrees) | 8 | 8 | 150 | -- | 89 |
| Voxel size (mm<sup>3</sup>) | 1x1x1 | 0.9x0.9x1 | 1.1x0.9x3 | 2x2x2 | 3x3x3.5 |
| FoV read (mm) | 192 | 240 | 220 | 192 | 192 |
| Fov phase (%) | 90.6 | 81.3 | 100 | 100 | 100 |
| Base Resolution | 192 | 256 | 256 | 96 | 64 |
| Phase Resolution (%) | 100 | 100 | 75 | 100 | 100 |
| Bandwidth (Hz/Px) | 130 | 130 | 201 | 1628 | 2368 |
| Orientation | Transversal | Transversal | Transversal | Transversal | Transversal |
| N volumes | -- | -- | -- | 60 directions + 5 b=0 | 180 |
| Other sequence-specific characteristics | -- | -- | -- | b-value = 1000 s/mm<sup>2</sup>; Echo spacing = 0.69 ms | Echo spacing = 0.49 ms; Eyes open |
| Acquisition time | 5 m 56 s | 9 m 8 s | 5 m 8 s | 11 m 11 s | 6 m 4 s |
| **Number of subjects** |  |  |  |  |  |
| N iPD | 119 | 118 | 100 | 114 | 117 |
| N PD-LRRK2 | 5 | 5 | 1 | 2 | 4 |
| N PD-GBA | 10 | 10 | 1 | 10 | 10 |
| N RBD | 74 | 74 | 68 | 74 | 74 |
| N RBD-GBA | 3 | 3 | 3 | 3 | 3 |
| N aGBA | 8 | 8 | 6 | 8 | 8 |
| N HC | 68 | 66 | 55 | 66 | 68 |
| N total | 287 | 284 | 234 | 277 | 284 |

Legend: iPD = idiopathic Parkinson's patients; PD-LRRK2 = Parkinson's patients with mutation of the LRRK2 gene; PD-GBA = Parkinson's patients with mutation of the GBA gene; RBD = patients with REM sleep behavior disorder (RBD); RBD-GBA = RBD patients with pathogenic mutation of the GBA gene; aLRRK2 = asymptomatic carriers of a pathogenic mutation of the LRRK2 gene; aGBA = asymptomatic carriers of a pathogenic mutation of the GBA gene; HC = healthy controls.

#### T1-weighted MRI

The T1w MPRAGE (Fig. 1a-1d) offers very good contrast across tissue classes: grey matter (GM), white matter (WM), and cerebrospinal fluid (CSF). It is primarily used to study GM structural macroscopic tissue in both cortical and subcortical regions. In Parkinson’s, cortical morphology in cognitively intact patients is generally reported to be normal or mildly altered, while impaired cognition and dementia in Parkinson’s have been found to be associated with more severe patterns of cortical atrophy^6 13-15^. Non-motor symptoms have been also associated with structural changes in specific related brain networks (for a review see ^6^). With this sequence we aimed to investigate potential differences in GM density across groups or relationships between GM and clinical variables, as well as taking into account the effect of structural changes when analysing functional MRI data.

**Figure 1.**
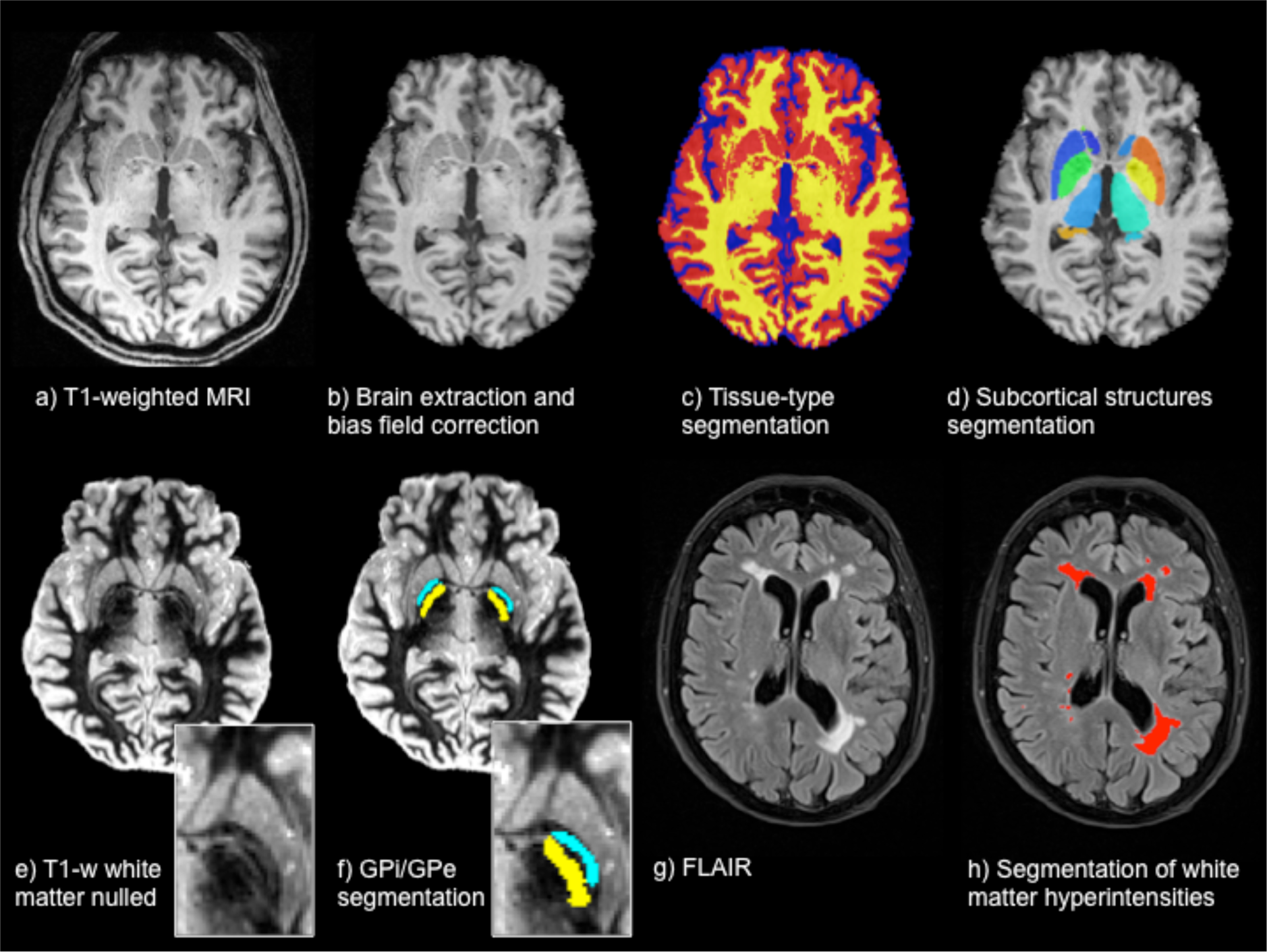
Structural sequences and related processing. T1weighted MRI (a) is brain extracted and bias field corrected (b) to perform tissue-type segmentation (GM in red, WM in yellow, CSF in blue) (c) and subcortical structures segmentation (d). T1-weighted white matter nulled (e) allows better contrast in the subcortical structures, which allows, for example, the segmentation of the Globus Pallidus (zoom) into its internal (GPi, yellow) and external (GPe, light blue) portions (f). FLAIR images (g) are used to detect and quantify white matter hyperintensities (red) (h).

The fsl_anat pipeline was used to perform brain-extraction, bias-field correction and 3-class (GM, WM, CSF) tissue-type segmentation with FAST^16^. The resulting images were then used to perform voxel-based morphometry analyses^17^ and to generate voxel-wise confound regressors for resting state functional MRI statistical analyses. Subcortical structure segmentation was performed with FIRST^18^, obtaining 3D meshes for each structure for each subject (used to perform vertex analysis), as well as volumetric images used to calculate volumes or as masks to extract average values from other modalities.

T1-weighted images with suppressed white matter signal (T1 WM nulled) show enhanced contrast across the basal ganglia structures (Fig.1e). This allows the detection of, for example, the boundary between the internal and external globus pallidus and a finer segmentation of this structure using MIST^19^ (Fig.1f).

#### T2-weighted Fluid-attenuated inversion recovery (FLAIR)

This sequence is commonly used to detect white matter abnormalities like leukoencephalopathies, demyelinating diseases and abnormalities of vascular origin. Regarding vascular pathology, it is used to detect white matter hyperintensities (WMH) (Fig. 1g,1h). With this sequence we wanted to characterise the amount and distribution of WMH in the cohort, look for differences across groups and assess the possible relationship with cardiovascular risk.

FLAIR images were brain extracted and bias field corrected using FAST^16^. WMHs were then automatically segmented on FLAIR images with BIANCA^20^, a supervised segmentation tool which assigns to each voxel a probability of being a lesion, based on their intensity in FLAIR and T1 and their location (details and training dataset are online^21^). The probabilistic output was thresholded and restricted to the voxels located within a mask created from the T1-weighted scans (using the command make_bianca_mask), which excluded the cortex and subcortical structures. The total WMH volume was then calculated for each scan.

#### Diffusion-weighted MRI (dMRI)

Diffusion MRI is used to study the microstructure of brain WM *in vivo*. Also, it serves to virtually reconstruct putative WM tracts. The basis of dMRI is measurement of the random motion of water molecules (diffusion), which has a preferential orientation in the WM (i.e. is less restricted along than across the axons). It follows that the preferential direction of water diffusion is related to fibre orientation. The amount of diffusion directionality and restriction can inform about the microstructural environment of a voxel under study.

Diffusion-weighted images were acquired along 60 isotropically distributed diffusion directions (b-value of 1000 s/mm^2^). Five additional images were acquired without diffusion weighting (b=0 s/mm^2^). B0 inhomogeneity for diffusion imaging was measured using a dual-echo GRE sequence and the resulting phase and magnitude images were processed to produce field maps for correction of inhomogeneity-induced distortions.

Correction for b0-associated and eddy current-related distortion, as well as participant’s movement, were performed using EDDY^22^. EDDY uses a generative probabilistic model to estimate inter- and intra-volume movements, displacements caused by field inhomogeneity, and distortions caused by eddy currents induced by the diffusion gradients. Additionally, automatic artefact rejection replaces slice drop-outs with model estimates^23 24^. The resultant 4D diffusion data were then fed into dtifit, which fits a diffusion tensor model at each voxel^25 26^ and generates maps of tensor-derived measures to assess WM microstructural integrity: fractional anisotropy (FA), mean diffusivity (MD), axial diffusivity (AD) and radial diffusivity (RD). Finally, we ran bedpostX to produce fibre orientation estimates and their respective uncertainties. This model can be used to perform probabilistic tractography for reconstructing WM pathways and assess their properties such as structural connectivity.

#### Resting-state functional MRI (rfMRI)

rfMRI is used to investigate brain function without requiring the subject to undertake a specific task. Although potentially harder to interpret than task fMRI, rfMRI is not affected by subject’s performance or compliance and allows to study different resting state networks, i.e. sets of brain regions sharing a common time-course of spontaneous fluctuations that have been associated with specific brain functions^27^. Among those, the basal ganglia network^28^ (supplementary Figure S1) is of particular interest for the OPDC-MRI cohort. In 28 iPD patients we also repeated the rfMRI sequence in the “on-state” 60–90 min after taking their own dopaminergic medication^11^.

Firstly, images were motion corrected with MCFLIRT and the six rigid-body parameter time series extracted for each subject were used for subsequent cleaning. Mean relative displacement was also calculated to potentially exclude subjects with excessive motion and as possible confound metric in further analyses. Images were brain extracted, corrected for B0 inhomogeneities using field maps, spatially smoothed using a Gaussian kernel of FWHM of 6 mm, and temporally filtered using a high-pass filtering of 150 s. Single-subject probabilistic independent component analysis (ICA) was then performed with MELODIC^29^ with automated dimensionality estimation, followed by automatic component classification with FSL-FIX^30 31^ to identify and regress out the contribution of the artefactual components reflecting non-neuronal fluctuations (FIX training dataset available online^21^). The pre-processed functional data were registered to the individual’s structural scan and standard space images using FLIRT and FNIRT^32 33^, using boundary-based-registration. Single-subject resting state networks were derived with dual regression^34^ and compared across subjects. The spatial maps used as spatial regressors in dual regression can be derived either from group-level ICA on the study-specific data^11^ or using an external template (for more details regarding this choice see ^35^). Supplementary Figure S1 shows the basal ganglia network map, part of the template used for the analyses. The full templates used for the studies published so far with data from the OPDC cohort are available online^21^.

Regarding non-core sequences acquired in the OPDC-MRI sub-study, Table 3 shows the main acquisition parameters and the number of available subjects per group for two sequences introduced in 2016, which showed particularly promising results. In the Supplementary material we provide more information on the other sequences experimented in this study, which include quantitative T1 and T2 mapping, diffusion weighted imaging of the substantia nigra and multi-echo T2*-weighted images of the substantia nigra.

**Table 3.**
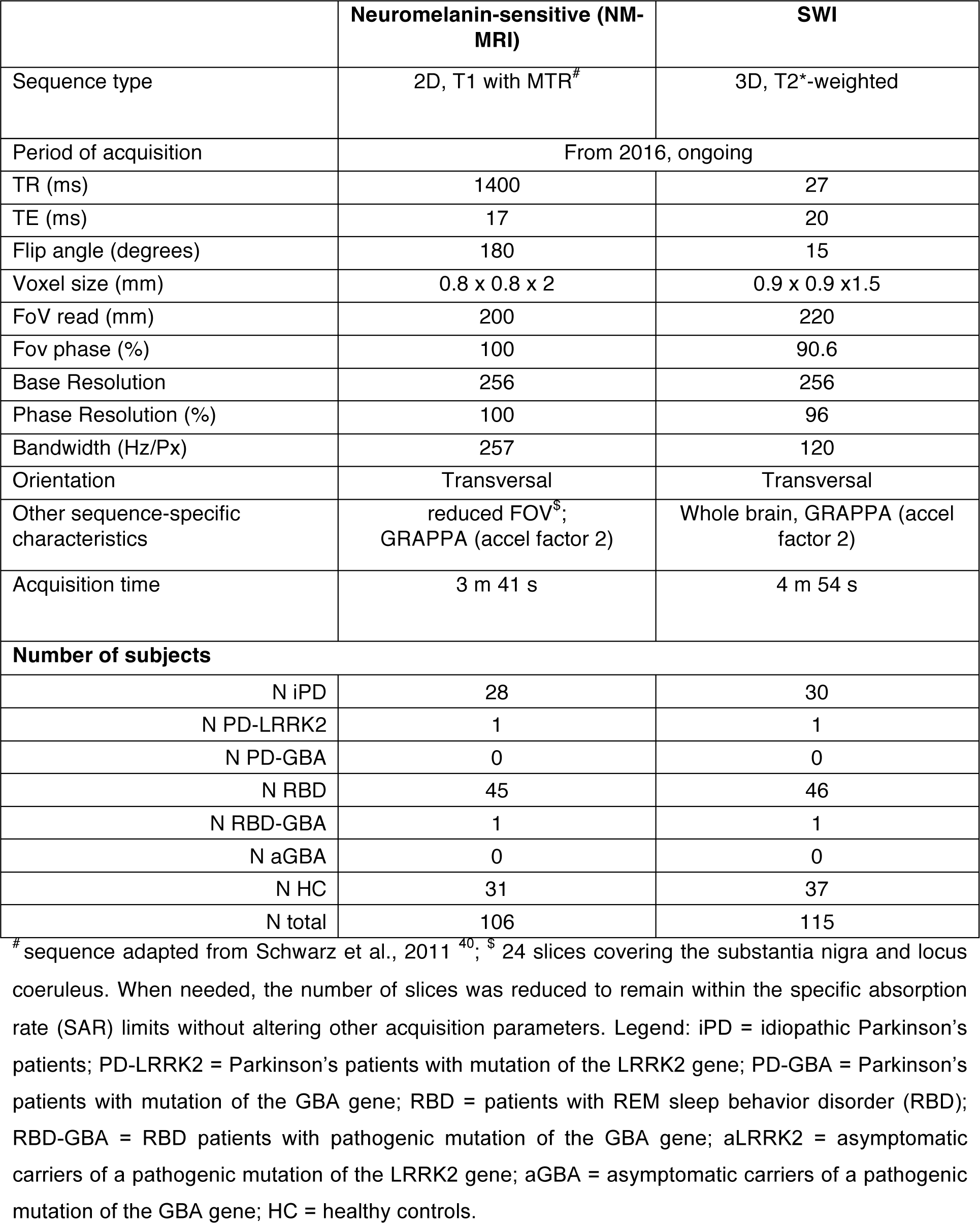
MRI experimental sequences: parameters used in the study and number of available datasets for each modality. Other experimental sequences are listed in the Supplementary material.

#### Neuromelanin-sensitive MRI (NM-MRI)

The monaminergic neurons in the substantia nigra (SN) and locus coeruleus (LC) are rich in neuromelanin, a dark pigment that gives these structures their distinct colour. Neuromelanin is detectable with MRI as a hyperintense signal using modified T1-weighted sequences^36 37^, which exploit the paramagnetic properties of the pigment, due to its iron content. According to pathological studies in Parkinson’s^38^, the SN and LC are affected early in the neurodegeneration process^39^, making them an interesting target for the development of neuroimaging biomarkers. Studies using NM-MRI found a reduction of the hyperintense signal on NM-MRI in SN and/or LC in patients in patients with established Parkinson’s^40 41^ and RBD^42-44^. These promising findings drove the inclusion of this sequence in our protocol.

NM-MRI images acquired in OPDC-MRI were bias field corrected using FAST^16^ and the transformations to the individual’s structural scan (T1w) and standard space were calculated using FLIRT and FNIRT^32 33^. We also defined two reference regions of interest (ROIs) in MNI space (one for SN and one for LC) and used the average intensities within the ROI (registered in individual subject space) as normalization factors in subsequent analyses.

To also extract quantitative information on these images, we developed a segmentation method to automatically quantify the hyperintense signal in SN and LC (Figure 2a and 2b, preliminary results in ^45^). The analysis on the whole sample is currently ongoing and support data related to the analyses (e.g. reference ROIs) will be available online^21^.

**Figure 2.**
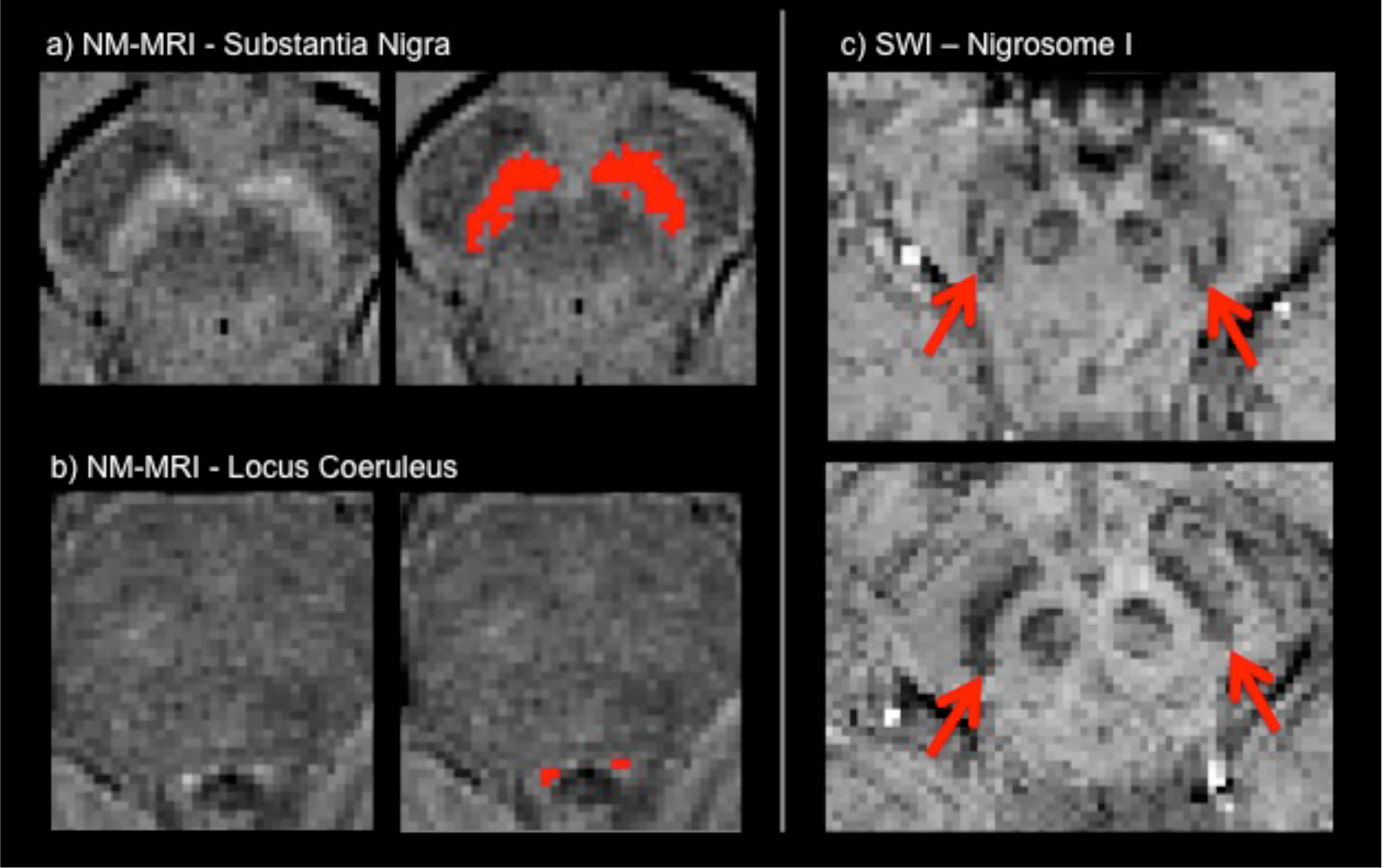
Novel sequences: Neuromelanin-sensitive (NM-MRI, left) and susceptibility weighted imaging (SWI, right). Examples of segmentation of (a) the substantia nigra and (b) the locus coeruleus as hyperintense areas on NM-MRI. (c) Examples of the presence (top) absence (bottom) of the ‘swallow tail’ sign on SWI.

#### T2*-weighted images / susceptibility weighted imaging (SWI)

Susceptibility-weighted imaging (SWI) uses tissue magnetic susceptibility differences to enhance contrast in MRI. This is achieved by using the phase image in addition to the magnitude of T2*-weighted images. The phase image contains information about local susceptibility changes between tissues, which can be useful in measuring iron content. There are numerous neurologic disorders that can benefit from a sensitive method that monitors the amount of iron in the brain, whether in the form of deoxyhemoglobin, ferritin, or hemosiderin^46^. In Parkinson’s, SWI has recently emerged as a promising sequence for evaluating the integrity of the substantia nigra^47^. In healthy subjects, the dorsolateral SN shows an area of signal hyperintensity, corresponding to nigrosome-I. This feature, described as a ‘swallow tail’ appearance^48^, is lost in Parkinson’s, since nigrosome-I is affected early by synuclein degeneration. Given the promising evidence for the ‘swallow tail’ to be a candidate biomarker for Parkinson’s, we included SWI in our protocol.

The following pre-processing was applied to the T2* images to obtain the final SWI image: macroscopic phase artefacts removal was performed by high-pass filtering the phase images using a 50×50 FWHM window in Fourier space (window size selected empirically to suppress artefacts in the midbrain caused by nearby aerated structures). Then, paramagnetic phase components only were taken to the 4th power and multiplied with the magnitude images.

The presence/absence of the dorsal nigral hyperintensity, the ‘swallow tail’ sign, was visually rated for each subject (Figure 2c). We also calculated the transformations from the individual’s SWI to structural scan (T1w) and standard space using FLIRT and FNIRT^32 33^ to perform group comparisons. To extract quantitative information on these images, we also developed a method to automatically quantify the nigrosome-I hyperintense signal^49^. The analysis on the whole sample is currently ongoing and support data related to the analyses will be available online^21^.

### Findings to date

Regarding brain structure, we found no differences between early iPD patients and controls using relatively standard analyses of whole-brain GM volume and overall regional volumes. A subtle abnormality in the shape of the right pallidus was detected, and corresponded to differences in connecting WM pathways. However, the subtle nature of these changes makes it unlikely that morphometric analysis alone will be useful for early diagnosis of Parkinson’s^50^.

We found evidence of alterations in microstructural integrity in the prefrontal cortex that correlate with performance on executive function tests^51^. This was investigated using cortical measurements of macro- and microstructure and performing multi-modal linked ICA on structural, quantitative T1 and dMRI in early Parkinson’s. Although these patients were cognitively intact, there were significant differences between Parkinson’s and controls in a fronto-parietal component primarily driven by altered cortical diffusion (FA and MD). These were related to cognitive performance. Intriguingly, the frontal areas involved match the distribution of impairment in striatal dopaminergic projections reported previously. A study in an independent patient group found a similar association between frontal WM integrity and cognition, corroborating the idea of early frontal microstructural involvement^52^.

Brain function, as assayed with resting fMRI yielded more substantial differences, with basal ganglia functional connectivity (BGFC) reduced in early iPD and increased upon administration of dopaminergic medication^11^. This alteration was not found in Alzheimer’s^53^, providing some confidence that the effect is pathology specific. A similar BGFC reduction to iPD was found in RBD^54^ and was replicated subsequently, although with smaller effect size, in a larger sample of this cohort (https://identifiers.org/neurovault.collection:5686). To test whether a link between BGFC and dopamine-related function was present in healthy aging, we conducted a multivariate analysis in a large population sample of healthy controls. We found an age-related and sex-dependent decline of connectivity, but no unique dopamine-related function seemed to have a link with BGFC beyond those detectable in and linearly correlated with healthy aging^55^. Our measure of BGFC was reproducible across different analysis settings^35^; however, these group differences have diminished with increasing sample size and have not been replicated on a different Parkinson’s population so far (unpublished). At the single subject-level, the discriminatory power of BGFC increased when using a more sophisticated supervised learning algorithm^56^, but further investigation is needed to assess the potential of rfMRI as a clinical biomarker.

Preliminary analyses of the novel sequences showed promising results. Measures of substantia nigra and locus coeruleus volumes extracted from NM-MRI were found to be reduced in RBD patients compared with controls (preliminary results in^45^). We also found a decrease in NM in the substantia nigra in Parkinson’s with respect to controls, especially in Parkinson’s with RBD, while the locus coeruleus seems more affected in RBD, in line with its role in REM sleep regulation.

Analysis of SWI images showed progressive reduction of the nigrosome-1 signal intensity (swallow-tail sign) from HC to RBD to manifest Parkinson’s in our cross-sectional sample. Preliminary results in our RBD imaging cohort show that 28% of patients have pathological nigrosome imaging, defined as absence of the dorsal nigral hyperintensity that represents nigrosome-1. Intriguingly, these patients also have reduced dopamine transporter binding in the striatum, suggesting that nigral SWI may be able to identify individuals with dopaminergic decline. This MRI method may have the potential to enrich cohorts for future neuroprotective trials in RBD, where participants with a high likelihood of conversion to motor Parkinson’s are sought to achieve clinical endpoints in a manageable timeframe^49^. However, longitudinal follow up to determine the true predictive value of SWI will be needed to confirm this.

#### Long-term follow-up

These baseline data already represent a rich source of data from a deeply phenotyped cohort. A key aspect of the OPDC-MRI cohort, however, is the longitudinal follow-up, which is ongoing. Clinical longitudinal data are acquired in the Discovery cohort every 18 months, and therefore, we will be able to use them to relate baseline imaging with clinical progression. This will potentially allow us to stratify patients and at-risk individuals and predict their progression. Information about conversion to Parkinson’s of at-risk individuals will also be available, providing the ultimate validation of potential biomarkers. In 2015 we also commenced the acquisition of longitudinal follow-up MRI after 5 years from baseline for Parkinson’s and HC, and after 2.5-3 years for RBD. This difference in time after baseline was chosen because we predict that a number of the RBD patients will convert to Parkinson’s, so we aim to collect multiple data points prior to conversion. In this way we hope to assess their trajectory during the prodromal phase and capture the associated brain changes.

### Strengths and limitations of this study

The main strength of this cohort is the collection of high quality 3T MRI data in a very well-phenotyped longitudinal cohort of Parkinson’s and at-risk individuals. The RBD dataset is one of the biggest brain MRI datasets available for this population of at-risk individuals.

Inevitably, a limitation is due to the trade-off between depth of the phenotyping and size of the cohort. OPDC-MRI phenotyping is deep and longitudinal, however the size of the cohort is relatively modest, particularly when compared with population-level cohort studies.

We are also aware of the inevitable selection bias for this cohort. We recruited people already participating in OPDC Discovery cohort and therefore our cohort reflects the demographic of the catchment area. Moreover, our participants are those who were already participating in a research study and declared they were happy to be contacted for imaging (self-selection). As a result, our cohort cannot fully capture the variability of Parkinson’s and RBD patients (e.g. patients with severe motor dysfunction may be less able to travel and tolerate an MRI scan).

Another key strength of the OPDC-MRI cohort is the imaging protocol. All MRI data were acquired using the same MRI scanner, which is quite unique for a study of this duration. In this way all baseline data, as well as longitudinal MRI data will be truly comparable, with no effect due to scanner and/or protocol change. The protocol includes both standard sequences, which are also comparable with other studies, as well as more experimental sequences acquired to investigate study-specific research questions.

We incorporated a trade-off between exploiting the latest techniques available in MRI and continuity of protocol throughout the study. While we could not use the most up-to-date sequences (e.g. our EPI images - dMRI and rfMRI - are not acquired with multiband acceleration), we managed to reach a balance by fixing the core sequences to maintain continuity, while changing the experimental sequences as the field evolved to include promising techniques as they became available.

As detailed in the previous section, the cohort will be enriched by collecting longitudinal follow-up MRI data. The challenge will be to be able to get data from those patients who have progressed quickly and will be in a more severe Parkinson’s stage. While they may still be able to undergo a telephone interview or clinical assessment, they may not be willing or able to tolerate an MRI scan.

As described in more details in the “Collaboration and data sharing” section below, we have made statistical maps of our results publicly available, as well as support data relative to the analyses. The data presented here are also available to request (details below).

### Collaboration and data sharing

Details about collaborating with OPDC can be found at https://www.opdc.ox.ac.uk/external-collaborations; OPDC is part of the CENTRE-PD twinning project (https://www.centre-pd.lu), we have ongoing international collaborations and are open for new proposals.

The data presented in this work (baseline imaging, demographics and clinical variables) will be available through the Dementias Platform UK (https://portal.dementiasplatform.uk), where data can be accessed by submitting a study proposal. Please note that longitudinal data will become available at a later stage.

Statistical maps are available on NeuroVault^57^ for the following publications:

^11^ https://neurovault.org/collections/2694/;

^53^ https://identifiers.org/neurovault.collection:5448;

^54^ (Results relative to a replication of the original study on an increased sample): https://identifiers.org/neurovault.collection:5686;

^35^ https://identifiers.org/neurovault.collection:2953

^55^ https://identifiers.org/neurovault.collection:2681.

Other types of support data related to the analyses are available online (https://ora.ox.ac.uk/objects/uuid:8200af66-f438-4a7b-ad14-e8b032f0a9e7)^21^ and the repository will keep being populated as the analyses progress.

## Data Availability

The data presented in this work will be available through the Dementias Platform UK, where data can be accessed by submitting a study proposal. Statistical maps are available on NeuroVault.org (links in manuscript) and support data related to the analyses are available on ORA Data.

https://portal.dementiasplatform.uk

https://ora.ox.ac.uk/objects/uuid:8200af66-f438-4a7b-ad14-e8b032f0a9e7

## Acknowledgments

The work was supported by the Monument Trust Discovery Award from Parkinson’s UK (J-1403) and by the Wellcome Centre for Integrative Neuroimaging, the MRC Dementias Platform UK, the National Institute for Health Research (NIHR) Oxford Biomedical Research Centre (BRC), and the NIHR Oxford Health BRC (a partnership between Oxford Health NHS Foundation Trust and the University of Oxford). The views expressed are those of the authors and not necessarily those of the NHS, the NIHR or the Department of Health.

JCK acknowledges support from the NIHR Oxford Health Clinical Research Facility. MR received funding support from a NIHR Academic Clinical Lectureship and a NIHR Oxford BRC Doctoral Training Fellowship. TRB received funding support from a Wellcome Trust Doctoral Training Fellowship, and a Biomedical Research Council Career Development Fellowship. FB received funding support from National Institute for Health Research.

The authors would like to acknowledge all the participants and their families for participating in the study. They also thank Tim Quinnell, Oliver Bandmann, Gary Dennis, Zenobia Zaiwalla and Graham Lennox for patients recruitment and in-clinic data collection. They are thankful to the staff of the Oxford Centre for Magnetic Resonance (OCMR), in particular Jane Francis, Kathryn Lacey, Rebecca Mills and Steven Knight, and to Amandine Louvel and Katie Ahmed for administering the cohort.

## Footnotes

### Author Contributions

LG had a major role in data acquisition and analysis, interpreted the data and drafted the manuscript for intellectual content.

JCK had a major role in data acquisition and analysis, interpreted the data and contributed to major revisions of the manuscript for intellectual content.

KSK, RALM, MR and TRB had a major role in data acquisition and analysis, interpreted the data and revised the manuscript for intellectual content.

ML, SE, FB, had a major role in data analysis and revised the manuscript for intellectual content.

MC and JR had a major role in data acquisition and revised the manuscript for intellectual content.

RWM, MTU, and CEM designed and conceptualised the study, interpreted the data and revised the manuscript for intellectual content.

All authors reviewed, critically revised and approved the manuscript.

### Conflicts of interest

Any conflict of interest is detailed in the authors’ disclosure forms, included in the submission.

### Data Sharing Statement

The data presented in this work will be available through the Dementias Platform UK (https://portal.dementiasplatform.uk), where data can be accessed by submitting a study proposal. Statistical maps are available on NeuroVault.org and support data related to the analyses are available online (https://ora.ox.ac.uk/objects/uuid:8200af66-f438-4a7b-ad14-e8b032f0a9e7).

## Notes

### Competing Interest Statement

MTH reports grants from Parkinson's UK Monument Discovery Award during the conduct of the study; other from Biogen Digital and Roche Prodromal Advisory Boards, outside the submitted work. Other authors have nothing to declare.

### Author Declarations

All relevant ethical guidelines have been followed and any necessary IRB and/or ethics committee approvals have been obtained.

Any clinical trials involved have been registered with an ICMJE-approved registry such as ClinicalTrials.gov and the trial ID is included in the manuscript.

